# Stress-induced changes in inflammatory and atherosclerosis markers during official basketball games in professional coaches

**DOI:** 10.1101/2021.09.12.21263441

**Authors:** Athina Zerva, Marianna Chronaki, Andrea Paola Rojas Gil, Nikolaos Paschalidis, Panagiotis Andriopoulos, Maria Tsironi

**Affiliations:** Faculty of Human Movement and Quality of Life Sciences, Department of Sports Organization and Management, University of Peloponnese, Sparta, Greece; Faculty of Human Movement and Quality of Life Sciences, Department of Nursing, University of Peloponnese, Orthias Artemidos and Plateon, 23100, Sparta, Greece; Biomedical Research Foundation, Academy of Athens, 4 Soranou Efessiou str., 11527, Athens, Greece

**Keywords:** stress, professional basketball coaches, basketball, immune system, inflammation, atherosclerosis, cytokines, cortisol, risk factors

## Abstract

**Objective:** Top-level competitive sports coaches repeatedly cope with situations of acute stress in order to succeed and manage high team performance. Occupational stress-induced biochemical and immune system markers are not well studied for this specific group of people. The purpose of this study was to evaluate stress-induced alterations of inflammatory markers and atherosclerosis risk factors during an official basketball game in top-level professional basketball coaches (head and assistant).

**Methods:** Blood samples and vital signs were obtained from 27 healthy coaches (Greek A1 Men’s National Basketball League), 30 minutes before and 30 minutes after the games. We performed a full blood count and measured inflammatory cytokines, atherosclerosis markers and cortisol levels. Data were statistically analysed using two-tailed paired and independent samples t-tests and Pearson’s Correlations.

**Results:** Post-game neutrophils (NEU) and apolipoprotein B (ApoB) levels were significantly increased while lymphocytes (LYM) were significantly decreased in comparison to pre-game values. Blood pressure (systolic and diastolic) levels were considered as a pre-hypertension state at both measurements. We found significant alterations between head and assistant coaches in diastolic blood pressure and cortisol levels after the game. Cortisol was negatively correlated to inflammatory cytokine levels and positively correlated with ApoB levels.

**Conclusions:** Game-induced acute psychological stress initiates an aseptic inflammatory response in top-level professional coaches and can be related to the atherosclerosis pathways posing as an acute as well as chronic health threat for top-level coaches who have to deal with long periods of stressful working conditions.

**SUMMARY BOX:** *What are the new findings?:*

- Top-level basketball games induce an aseptic inflammatory response in professional basketball coaches mainly manifested by increased levels of peripheral blood neutrophils
- The response to game conditions also included elevated levels of ApoB after the game that were positively correlated with cortisol levels
- Stressful game conditions over a long season of games could pose as a serious health issue for professional coaches
- Health monitoring for professional coaches in top-level competitive sports such as basketball should be implemented more often with particular focus on inflammatory and atherosclerosis markers

## INTRODUCTION

The profession of coaching is distinguished for its uniqueness, due to the complexity of duties, the multiple roles and the high professional standards. In the age of intense sports management and marketing, professional sports have evolved to be “too much of a game as to be characterized as an industry and too much of an industry as to be characterized as a simple game”^1^. The implication of coaches in human resources management, their evaluation by many different persons implicated in teams’ events and the continuous effort for successful performance of the team are reported as some of the factors that may lead to severe stress situations^1 2^.Stress at work constitutes today a large area for research which is recognized worldwide as a serious threat for employees’ health^3^. Psychosocial stress originating from the daily working environment (occupational stress) is reported by the World Health Organization (WHO) as a risk factor of cardiovascular diseases while many studies have also shown that psychological stress induces increase of hormone levels associated with stress^4 5^, alterations of plasma lipid levels^4^ and also immune system factors^6^. Many studies have been conducted in the real working environment of the participants, on many different professions, using simultaneously descriptive and experimental methods, showed that the alterations provoked in biochemical indicators due to stress at work, cumber employees’ health^7-10^. To date, very few studies have addressed these issues for the profession of basketball coaching.

Each time the human body is encountered with stress factors, it attempts to achieve adaptation in the new condition through a number of physical and mental reactions, known as “*adaptive response*” that involves the participation of the neuroendocrine system (the Hypothalamic-Pituitary-Adrenal (HPA) axis – Cortisol) and the immune system^11^ 12. Brain hypothalamus produces the Corticotropin-Releasing Hormone (CRH) which stimulates the secretion of the Adrenocorticotropic Hormone (ACTH) from the pituitary gland. ACTH, through blood circulation, stimulates the production of cortisol by the adrenal glands^12^ triggering the increase of heart rate and blood pressure, peripheral vascular resistance, increased muscle tension and respiratory disorders^13^. Involvement of the immune system in response to stress is manifested through an inflammatory response which is otherwise a crucial response against infections. It is regulated by cells like neutrophils and macrophages and controlled by cytokines, released by these cells, such as IL-1, IL-6 and TNF-α^14^. Cytokines act as special chemical mediators including a variety of actions like the activation of endothelial cells which further attract and activate circulating monocytes and lymphocytes^14 15^. The participation of the immune system in response to stress could lead to a state of “low-grade” inflammation that ultimately could trigger adaptive immune responses of a chronic nature^16^.

Atherosclerosis is slowly evolving in the arterial tunica intima and tunica media while its effects lead to arterial stenosis, reduced blood flow in target-organs and formation of atheromatous plaques, prone to rupture. Smoking, dyslipidemia, diabetes mellitus, hypertension, obesity, metabolic syndrome (hypertension, dyslipidemia, insulin resistance/glucose intolerance/diabetes mellitus, abdominal obesity) and physical inactivity (absence of exercise) are reported as modifiable risk factors for atherosclerosis while current atheromatous lipid parameters include increased lipoprotein-a (Lp-a) and apolipoprotein B (Apo-B) levels, and reduced apolipoprotein A-I (ApoA-I) levels^17^.

In sports medicine, the “exercise stress” model has been widely studied. Using this model, several studies demonstrated that intense and prolonged exercise may provoke alterations in hormonal indicators and risk factors for atherosclerosis as well as mobilization of immune cells and activation of the inflammatory response^18-24^. Previous research on stress related effects in coaches has been mainly performed with the use of questionnaires, interviews and individual case-studies and focused on measurement of cortisol and cardiovascular function^25-27^. These studies involved the analysis of personal, social, demographical, environmental and working stress factors^28-31^. Indicative subjects examined in these studies were, salaries, occupational duties and responsibilities, tensions between working and family environments and risk of mental-emotional exhaustion (‘burn-out’ syndrome).

The main aim of our study was to measure inflammatory markers and atherosclerosis risk factors in the blood of professional coaches before and after official basketball games.

## MATERIALS AND METHODS

### Participants

In the present study, 27 professional basketball coaches (n=27, head coaches and assistant coaches) were included. The 27 coaches participated in the Greek Basket League’s official A1 Men’s Division games taking place in Greece, organized by the Hellenic Basketball Clubs Association (HEBA). For each participant, a brief medical history was taken, including information for anthropometric (height, weight) and demographic characteristics (sex, age). Written consent was obtained and records confidentiality was retained. 6 of the participants were smokers (10-20 cigarettes/day for a time period of 7-25 years).The main inclusion criterion was absence of infection for the last three weeks while the main exclusion criterion was steroid treatment and/or immunotherapy. Participants were not involved in the design, or conduct, or reporting, or dissemination plans of our research.

### Bioethics

The study was approved by the Bioethics Committee of the University of Peloponnese and was carried out in accordance with the Declaration of Helsinki (1989). The participants were informed about the aims and procedures of the study. A signed consent was obtained before the completion of the tests. The anonymity of the participants was assured.

### Blood collection and blood pressure measurements

In total, blood samples were collected before(without prior fasting) and after the game in eight respective official games of the Basket League between 13h30 and 21h00 (game times). Coaches underwent measurement of blood pressure-two measurements of systolic and diastolic arterial blood pressure were performed with a mean interval of 5 minutes, in sitting position-and subsequent blood sample collection (phlebotomy) 30 minutes prior and 30 minutes after the game.

### Lipid profile, leukocytes and cytokine assays

Blood samples were analyzed for lipid profile (cholesterol, triglycerides, HDL, LDL, Lpa, ApoA and ApoB) using the analyzer Medilyzer (Medicon, Hellas).Leukocyte, HGB and HCT levels were also measured. The complete blood count (CBC) was performed through blood analyzer (ABBOTT, USA). Inflammatory cytokines (IL-1β, IL-6, IL-8, IL-12p70, and TNF-α) were measured with multiplex method and Cytometric Bead Arrays (CBA) according to the manufacturer’s instructions (Human Inflammatory Cytokines Kit, BD Biosciences, San Jose, CA). Briefly, 50μl of the beads solution and 50μl of blood plasma from each participant were used. Samples were then incubated for 90 minutes protected from light. 1 ml of washing solution was added in each tube and centrifugation was performed at 200 x g for 5 minutes. Supernatants were cautiously removed and beads were re-suspended in 300μl of washing solution. Samples were analyzed with flow cytometry (FACSCalibur, BD Biosciences).

### Cortisol assays

Measurements of cortisol plasma levels were performed with Cortisol EIA Kit (competitive ELISA, Cayman Chemical Company, AnnArbor, Michigan) according to the manufacturer’s instructions. Results were measured using a plate reader (spectrophotometer) at wavelength 405-420nm.

### Statistical analysis

Statistical analysis was performed using Prism software (GraphPad Software, La Jolla, CA, USA) and SPSS 17.0 (IBM). All values were expressed as average means and statistical analysis was assessed by Student’s t-test for paired measurements (two-tailed paired t-test). Comparison between groups was performed using t-test for independent samples (independent samples t-test). Pearson’s correlation coefficients between cortisol values and cytokines values as well as Apo-B values before and after the game were also calculated. A probability of p< 0.05 was considered significant.

## RESULTS

### Stress induced changes in blood pressure and atherosclerosis related biochemical markers

Most of the participants in this study were holders of coaching diplomas with many years of experience of tenure in high level coaching in Greek and international professional league teams. The main characteristics of the sample are shown in table in Figure 1 (**Figure 1A**). We recorded age, height, weight and body mass indices (BMI) from all the participants. No significant difference was observed between the average values of blood pressure before and after the game (**Figure 1B**). However, a significant increase of post-game diastolic blood pressure was observed in head coaches as well as a significant difference between head and assistant coaches in regards to the change of diastolic blood pressure in terms of pre and post-game levels (**Figure 1C**). In particular, while the mean values of pre-game diastolic blood pressure did not differ for head and assistant coaches, post-game diastolic blood pressure levels of head coaches were significantly higher than in assistant coaches. No significant differences between the two groups were observed in relation to the systolic blood pressure changes before and after the game (data not shown).

**Figure 1.**
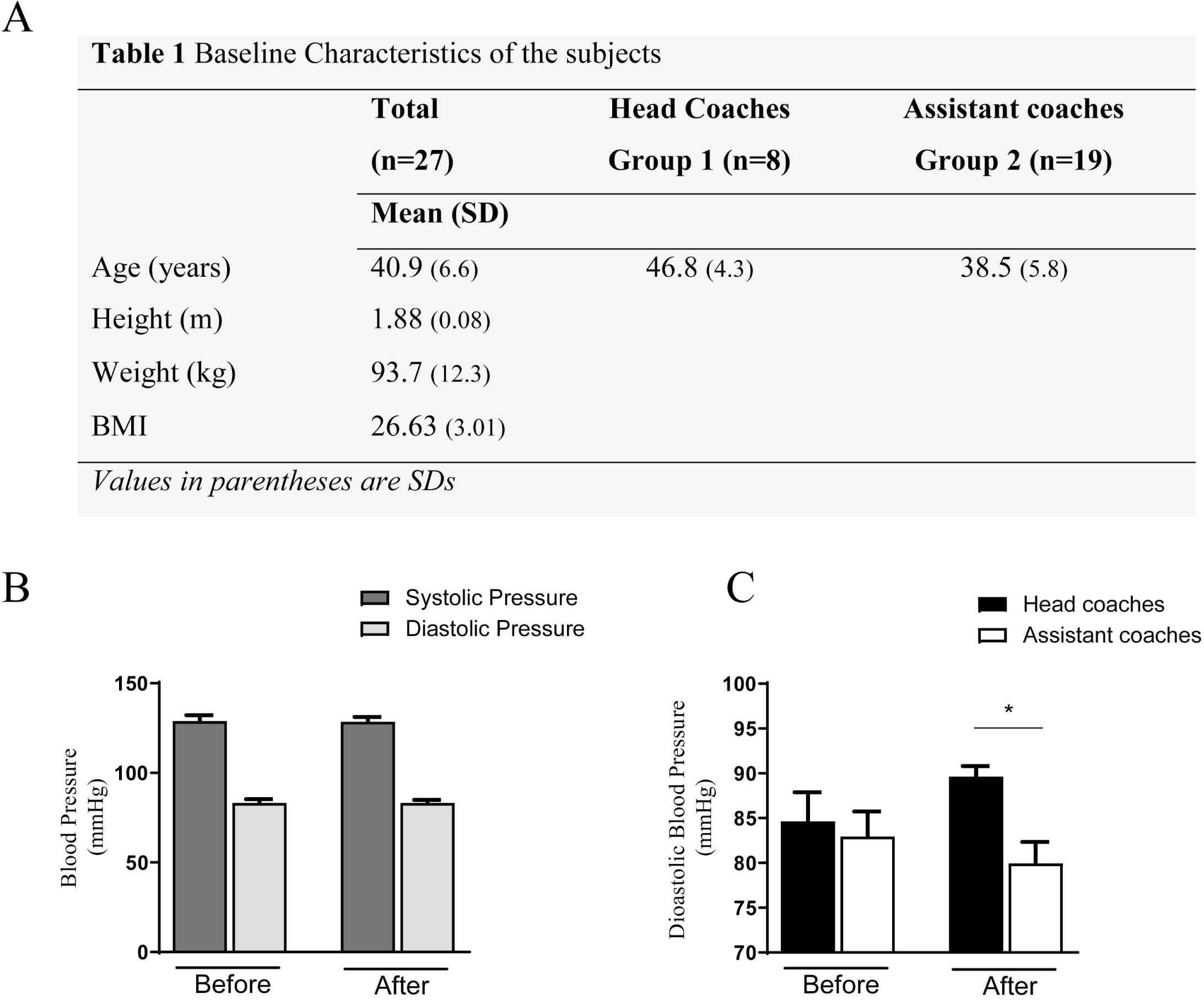
**(A)** Descriptive statistics of the mean age, height, weight and BMI (Values are given as mean ±Standard deviation) and (B) pre and post-game systolic and diastolic pressure values of participants. (C) Pre and post-game diastolic pressure levels for head (black bars) and assistant (white bars) coaches. Values are expressed as means ± Standard deviation. * p<0.05 (Students t-test).

In order to investigate possible game induced (stress factor) changes in biochemical markers related to atherosclerosis we measured blood levels of total Cholesterol (CHOL), Triglycerides (TRIGL), High Density Lipoprotein (HDL), Low Density Lipoprotein (LDL), Lipoprotein a (Lpa) and apolipoprotein AI and B (Apo AI, Apo B) (**Figure 2A**). We observed a statistically significant post-game decrease of total cholesterol and triglycerides. Most importantly, we also observed statistical significant decreased HDL, increased Apo B and. No statistically significant changes in Lpa and Apo A1 levels were observed.

**Figure 2.**
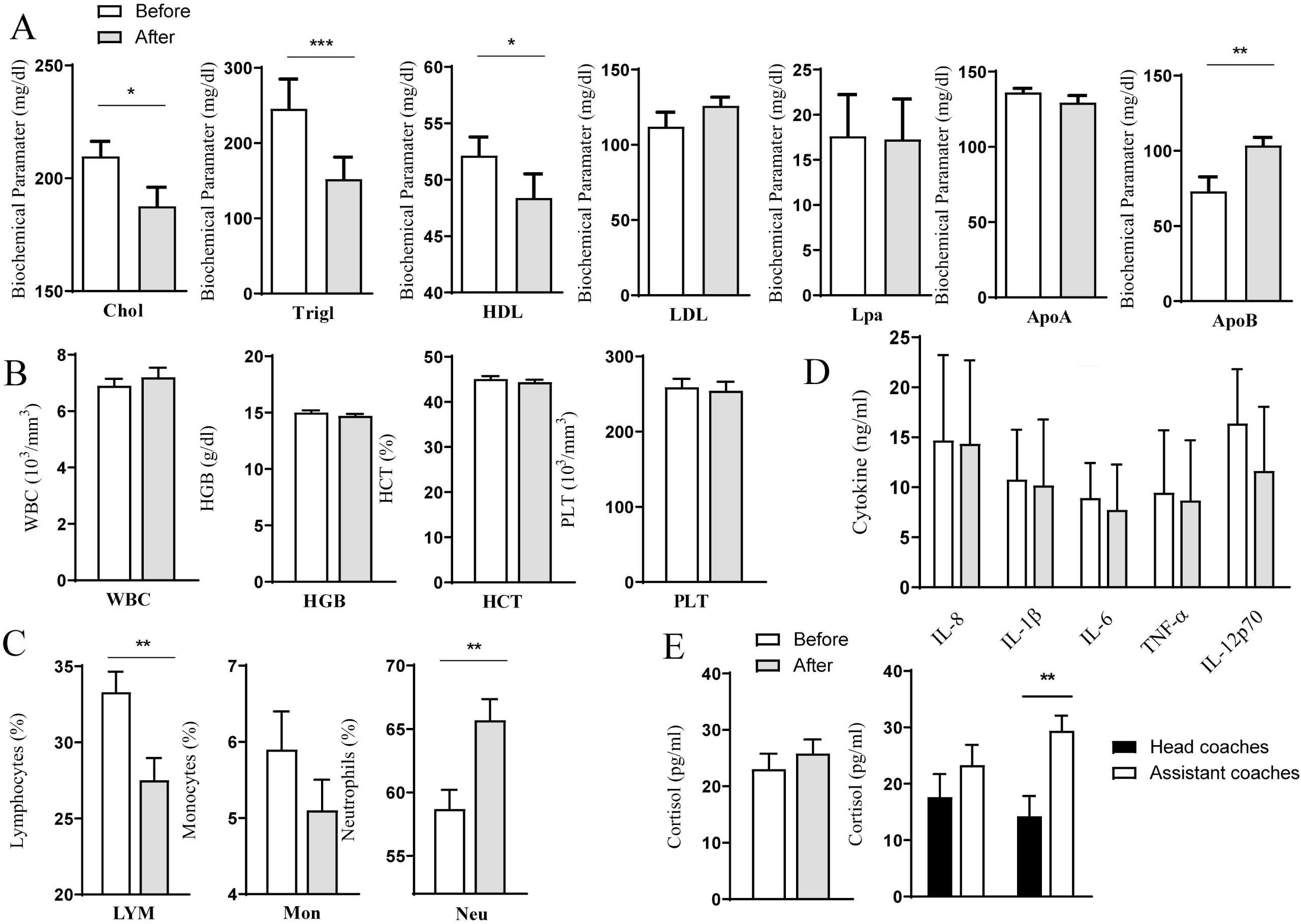
Pre and post-game biochemical parameters (A and B), immune cells (C), inflammatory cytokines (D) and cortisol (E) levels. Values are expressed as means ± Standard deviation. * p<0.05, **p<0.01 (Students t-test).

### Stress induced changes in inflammatory markers and leucocytes

We did not observe differences in white blood count (WBC), red blood count (RBC) platelets (PLT) and hematocrit (HCT) levels before and after the game (**Figure 2B**). However, a statistically significant decrease in lymphocytes (LYM) percentage was observed (pre-game mean value 33.3%, 27.5% post-game mean value) as well as increase in neutrophils (NEU) percentage mean value (pre-game 58.7% vs. post-game 67.7%) (**Figure 2C**). Although we observed decreased percentages of monocytes after the game, these differences were not statistically significant. We then measured the levels of IL-8, IL-1β, IL-6, IL-12p70 and TNF-α. We did not observe any significant changes in the values of these cytokines. Although the levels of some of these cytokines were reduced after the game, no significant alterations were measured (**Figure 2D**). It is known that stressful conditions can induce cortisol production. We measured cortisol levels before and after the game and we observed no differences for all subjects (**Figure 2E, left**). It is interesting to note that when the two groups (Head and assistant coaches) were analyzed separately we found that the group of head coaches had a relative decrease in cortisol levels after the game while on the contrary the group of assistant coaches had a relative increase (**Figure 2E, right**). Among these alteration we observed that total cumulative cortisol levels in assistant coaches group was statistically significantly higher than that of head coaches.

### Correlations between cortisol and inflammatory and atherosclerosis markers

Since cortisol is the main stress induced hormone that can affect inflammatory markers we analyzed the correlation of cortisol levels to the cytokines in all the subjects before and after the game. Cortisol values were negatively correlated to cytokines levels, with a stronger effect for the pre-game values (**Figure 3A**). More specifically, cortisol levels were correlated to IL-8, IL-1β, IL-6 and IL-12p70 concentrations, with a high significant correlation to IL-8 and IL-1β. (**Figure 3B**). Interestingly, post-game cytokine levels were strongly correlated to pre-game cortisol values and statistical significant differences were observed for IL-8, IL-1β, TNF-α and IL-12p70 (**Figure 4A and B**). Among all atherosclerosis related parameters, cortisol was positively correlated only to pre-game Apo-B values, but this was not confirmed for post-game values (**Figure 4C**).

**Figure 3.**
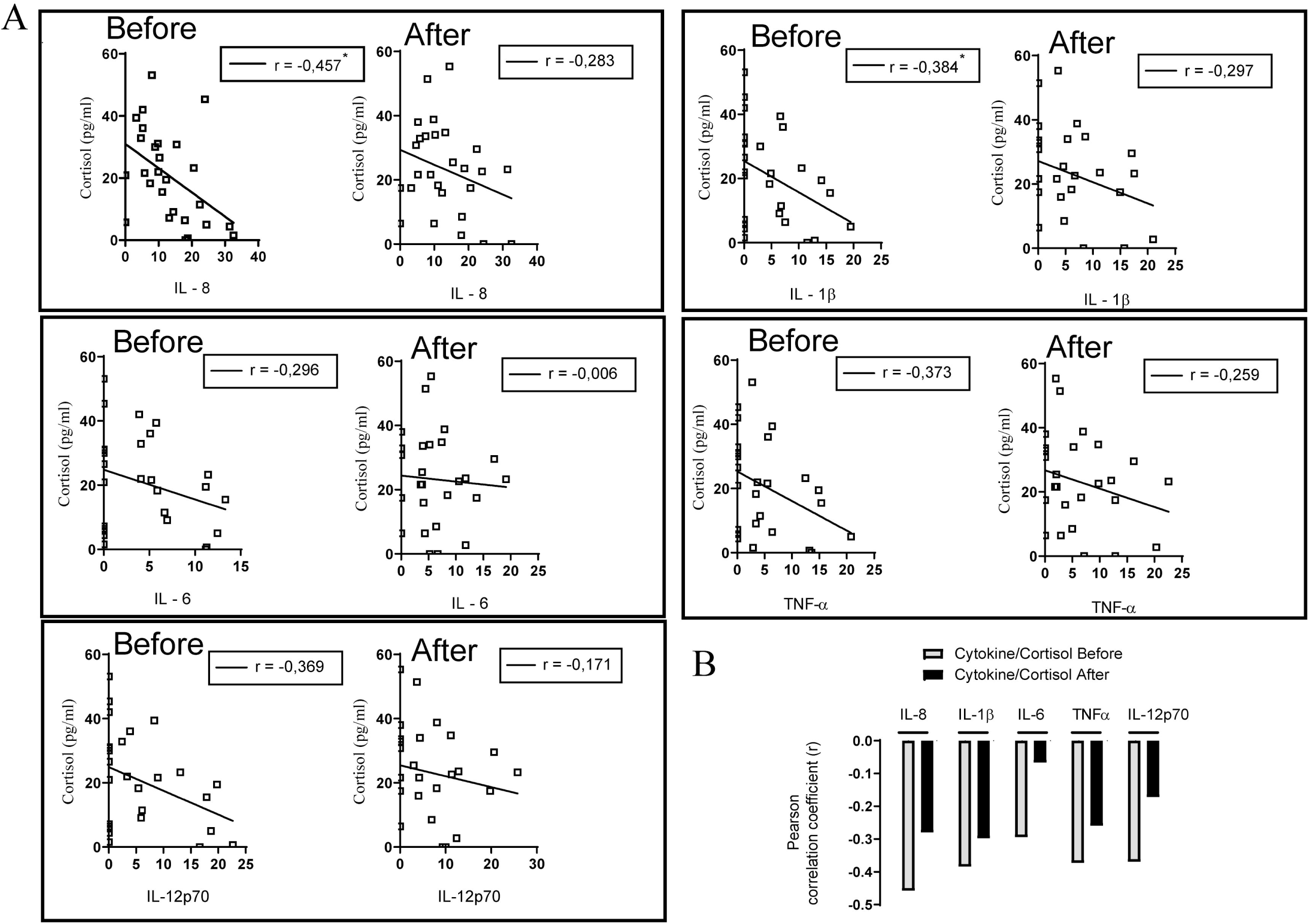
(A) Pre and post-game cortisol and Pre and post-game inflammatory cytokines correlation (B) cumulative values(r)*p<0.05(Students t-test).

**Figure 4.**
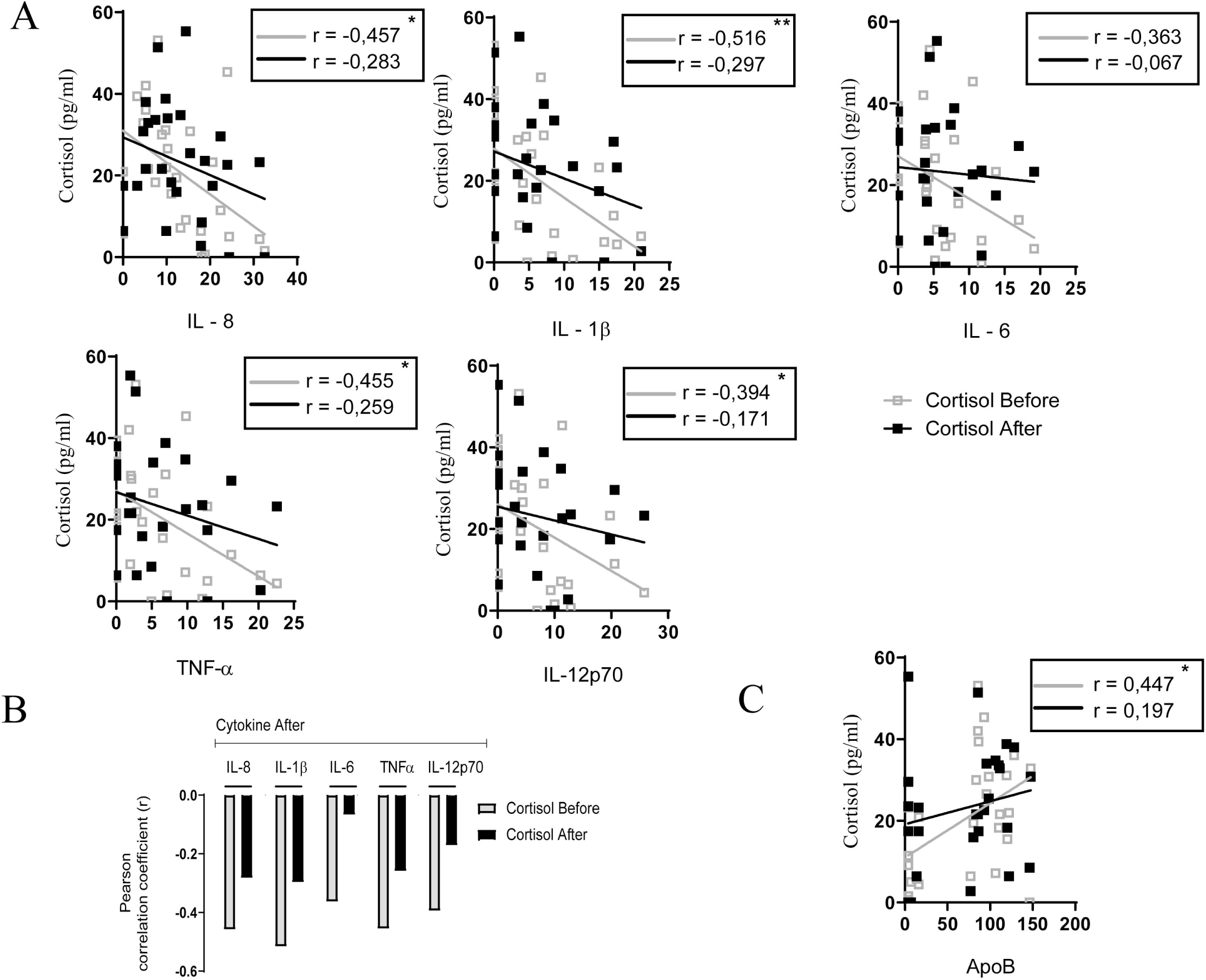
(A) Pre and post-game cortisol and post-game inflammatory cytokines correlation (B) cumulative values of r (Pearson correlation coefficient) for these correlations in all subjects. (C) Pre and post-game cortisol levels correlation to post game ApoB levels.*p<0.05, **p<0.01 (Students t-test).

## DISCUSSION

The purpose of this study was to examine the impact of an official high level basketball game on cardiovascular and endocrine systems of professional basketball coaches. Despite numerous studies examining immune-inflammatory and neuroendocrine responses induced by “exercise stress” in different athletes, the role of occupational induced stress and its effects to professional coaches has been neglected.

### Stress induced blood pressure

Although, no history of increased arterial blood pressure was noted in participants, some of them had slightly high blood pressure levels^32^. This was caused probably by sympathetic system stimulation (“flight or fight” system)^33^.This has been experimentally demonstrated by different studies, after exposure to acute psychological stress. Significant differences between blood pressure in head and assistant coaches before and after the game were observed and this could be attributed to the difference in the mean age between these two groups. Age is directly proportional to increases in blood pressure^34^. Furthermore, game conditions require higher physical and psychological effort from the head coaches (always in action and on the move, having the main control and the overall responsibility for the guidance of their teams).

### Effect of stress on the lipidemic profile

Acute stressful stimulation of short duration leads to an increase of lipid and lipoprotein levels^35-37^, although there are studies supporting that this increase is mainly attributed to plasma volume changes (hemoconcentration) ^2^. Previous studies have shown that occupational stress can affect the lipidemic profile which directly reflects on atherosclerosis predisposition^9^. In our study, a significant post game decrease in total cholesterol levels, triglycerides and High Density Lipoprotein (HDL) was observed, while Apolipoprotein-B (Apo-B) levels were increased. The decrease in triglycerides may be explained by basic metabolism mechanisms, while the decrease in HDL as well as the increase in Apo-B demonstrate an atherogenic lipidemic profile^38^. In support to our data, a large observational study (AMORIS, 68-month duration, 175.000 participants), showed that Apo-B was the most powerful and significant predictor of myocardial infarction^39^ and levels are not affected by non-fasting conditions^40^. Additionally, Apo-B values were increased in employees during periods of high workload in combination with their effort for achievements^9^. It is important to highlight that during intense physical stress due to prolonged exercise (e.g. ultra-marathon) an anti-atheromatic lipidemic profile has been observed^21 24^, whereas in our study, an acute psychological stress (official basketball game conditions) results to an increase of atheromatic risk factors levels.

### Effect of stress on immune system

Acute stress of short duration, such as professional basketball games in our study, can activate the immune system. In this study, elevated numbers of neutrophils (first line of defense of the immune system – innate arm) were observed after the game. The exact mechanism by which inflammation leads to elevated numbers of circulating neutrophils remains unknown^41^. Neutrophil increase was also observed in other studies during acute psychological stress ^14 41 42^. We also observed a post-game decrease in lymphocyte and monocyte levels that may be attributed to the proportional increase of neutrophils. As the game duration was two hours, these findings are in accordance another study which supports that acute stressful stimulation, of short duration (few minutes to hours), leads to an increase of lymphocytes and monocytes during the first 5-30 minutes, which is followed by their decrease within a time interval of 30 minutes to 4 hours -observed in our study^43^.

### Effect of stress on cortisol level

Regarding cortisol, the main hormone of the stress system, high levels of cortisol are observed 10-30 minutes after exposure to stressful conditions, returning back to normal after an hour ^44^. In our study pre and post-game cortisol values, for all participants appeared to be on a high range with an increasing tendency 30 minutes after the game. Interestingly when comparing the groups of head and assistant coaches, higher post-game cortisol values, almost double, were observed in assistant coaches group.

The tendency for increase of cortisol, observed after the game in all subjects, could be attributed to the larger group of assistant coaches (n=19) and the higher mean levels of cortisol they have presented. These results are in accordance with other experimental studies that explored psychological stress and distinguished the participants to high and low responsiveness to stress, depending on high and low cortisol concentrations respectively^5 45 46^.Additionally, the difference of age between head coaches and their assistants, the management of anger, tension and aggressive behavior in conjunction with the greater coaching experience that head coaches have, probably explains the reduced responsiveness of this group to game-induced stress, following the pattern of chronic stress. This phenomenon has also been described in lab experiments for cortisol levels measurements being under repetitive challenges and exposure to the same stressful impulse^45 47-50^.

Pre-game cortisol levels were negatively correlated to pre and post-game cytokine levels and these findings are in agreement with the study of Kunz-Ebrecht SR et al^51^. The negative correlation of cortisol and pro-inflammatory cytokines strengthens the hypothesis that cortisol might have an important and crucial role in the decrease of cytokine values after the game. These results are similar to other studies, since glucocorticoids (cortisol) suppress the expression of pro-inflammatory cytokines, such as IL-1, IL-2, IL-6, IL-8, IL-11, IL-12, TNF-α and upregulate the expression of anti-inflammatory cytokines (IL-10, IL-4)^52 53^.

Considering the physiology of the stress system, our study supports the induction of inflammation in basketball coaches, under the effect of the game as a stressful stimulant. Although white cells count remained almost the same after the game, differences observed on leukocytes subpopulations, as well as the high levels of cytokines in comparison to the general population values ^54^seem to be related to the activation of the inflammatory process^14 33^. The physical response to stress, mediated by the actions of cytokines is the activation of the central and sympathetic nervous systems, the activation of the HPA axis and the subsequent release of cortisol. Cortisol induces the negative feedback mechanism to the HPA axis but also regulates further immune responses^53^. Through catecholamines increase, circulating neutrophils may increase 4- or 5-fold, within a few hours hence provoking the initiation of the inflammatory response^33^, following the pattern of aseptic inflammation in cases of hemorrhage, trauma, hypoxia and treatment with glucocorticoids^55^. In this study, the cortisol secretion seems to have an important role on the increase of leukocytes and the decrease of the number of lymphocytes and monocytes in peripheral blood. The initial phase of the inflammatory response induced the redistribution of the leukocytes in the peripheral bloodstream. During the first minutes of the acute stressful response, the activation of the sympathetic system and the secretion of catecholamines and cortisol induce the sharp increase of leukocytes, and especially neutrophils, in the bloodstream^42 43^. During late stages of the stressful response and while stress conditions subsist up to a few hours or under conditions of severe psychological or physical stress, the initial increase of leukocytes (mainly neutrophils) after acute stress is followed by a decrease in their number due to the activation of the HPA axis. Additionally, the secretion of glucocorticoids is initiated leading to the sharp increase of leukocytes into the skin, the lungs, the gastrointestinal system, the urinary system, the mucosal system and the lymph nodes from the bloodstream^43 56 57^.

The activation of the HPA axis during stress conditions and the secretion of cortisol may also have an important role in the activation and the metabolism of lipids as well as CNS and HPA axis interaction which lead to an increase of catecholamines (adrenaline-noradrenaline)^4^. The statistically significant correlation of cortisol to ApoB, has been also discussed by a study of Sokolov and colleagues suggesting that hormones like insulin, cortisol and ACTH, affect the regulation of lipid metabolism ^58^. In the study of Nanjee and Miller, cortisol was positively correlated to LDL and the authors commented that this cortisol action on LDL metabolism is independent of Apo-B concentrations^59^.

## Conclusion

In conclusion, basketball game acts as an aseptic inflammation stimulus for professional coaches, triggering their stress system and up-regulating atherosclerotic factors. Taking in mind that professional coaches have to deal with a great number of games over a season and usually (unlike athletes) many years or decades of high level professional coaching practice, this poses as an immediate threat for them. It would be of great importance to further investigate and monitor immune and endocrine responses over the period of a full game season or multiple seasons, assuming that they are exposed to high levels of occupational stress. It would be also extremely useful to identify the initiation point of stress induction, any possible recovery period and whether these coaches have to deal with chronic stress symptoms throughout their career, lasting especially for high level ones for several decades.

## Data Availability

The data that support the findings of this study are available from the corresponding author upon reasonable request.

## AUTHOR CONTRIBUTIONS AND FUNDING

AZ and MT conceived and designed the study. AZ, MC and APRG assisted with recruitment of participants and performed sample collection. AZ and NP collected the data and conducted statistical analysis. All authors participated in data interpretation, manuscript preparation and approved the final manuscript. The authors received no financial support for the research conducted in this article.

## ACKNOWLEDGMENTS

We thank the Greek Basketball Coaches Association (SEPK), the Hellenic Basketball Clubs Association (HEBA) for the valuable assistance in the recruitment of participant in this study. We also thank the Hellenic Red Cross (Volunteers Samaritans, Rescuers and Lifeguards Corp) for assistance in blood collection and Apostolos Stergioulas for his assistance at the initial stages of the design of this study.

## HUMAN PARTICIPANTS AND ETHICS

The study was approved by the Bioethics Committee of the University of Peloponnese and was carried out in accordance with the Declaration of Helsinki (1989). The participants were informed about the aims and procedures of the study. A signed consent was obtained before the completion of the tests and records confidentiality was retained. The anonymity of the participants was assured. Participants were not involved in the design, or conduct, or reporting, or dissemination plans of our research.

